# Endoscopic Endonasal Surgery for Pituitary Adenomas with Cavernous Sinus Invasion: A Comprehensive Meta-Analysis of Efficacy, Remission Rates, Surgical Outcomes, and Complications

**DOI:** 10.1101/2025.08.31.25334779

**Authors:** Jaymin Pansuriya, Priyadarshini Panneerselvan Sundararajan, Sheikh Ramiz Ahmed, Sneha Baiju, Yashas Maragowdanahalli Somegowda, Yousra Merdjana, Arshi Wasim, Manav Kamleshkumar Patel, Saket Dineshkumar Prajapati, Rakhshanda Khan, Harshawardhan Dhanraj Ramteke, Manish Juneja

## Abstract

**Introduction:** Pituitary adenomas with cavernous sinus invasion represent a challenging subset of intracranial tumors. The presence of cavernous sinus invasion complicates surgical resection and increases the risk of recurrence and postoperative complications. Endoscopic endonasal surgery (EES) has emerged as a promising alternative to traditional microscopic transsphenoidal surgery, offering enhanced visualization and reduced collateral damage. This meta-analysis aims to evaluate the efficacy and safety of EES in the treatment of pituitary adenomas with cavernous sinus invasion, focusing on remission rates, gross total resection (GTR), recurrence rates, and complications.

**Methods:** A comprehensive literature search was conducted through PubMed, Scopus, Embase, and CENTRAL databases until August 2025, following PRISMA guidelines. Studies meeting eligibility criteria, including human studies published in English and involving adult patients with pituitary adenomas and cavernous sinus invasion treated with EES, were included. Data extraction focused on patient demographics, tumor characteristics, surgical outcomes, and complications. Statistical analysis was performed using a random-effects model, and subgroup analysis was conducted based on factors such as tumor size, Knosp grade, and hormonal subtype.

**Results:** A total of 27 studies involving 3,591 patients were included. The pooled remission rate was 60% (95% CI: 49% to 71%), with substantial variability across studies. The pooled residual tumor rate was 15% (95% CI: 11% to 19%), and the recurrence rate was 8% (95% CI: 6% to 11%). The incidence of cerebrospinal fluid (CSF) leaks was 9% (95% CI: 3% to 14%), while the rates of ICA injury and cranial nerve injury were extremely low (0.00% and 0.01%, respectively). Subgroup analyses revealed higher remission rates in macroadenomas (68%) compared to microadenomas (33%), and GH-secreting tumors showed higher endocrinological remission rates compared to ACTH-secreting tumors.

**Conclusion:** Endoscopic endonasal surgery demonstrates moderate efficacy in the treatment of pituitary adenomas with cavernous sinus invasion, with favorable remission and resection rates. However, substantial variability across studies emphasizes the need for further standardization of surgical techniques and patient selection criteria. The procedure is generally safe, with low rates of serious complications such as ICA injury and cranial nerve damage. Further prospective studies are needed to optimize patient management and evaluate long-term outcomes, including recurrence and quality of life.

## Introduction

Pituitary adenomas are among the most common intracranial tumors, accounting for approximately 10-25% of all primary brain neoplasms [1]. These tumors arise from the epithelial cells of the pituitary gland and can be classified based on their size and functional characteristics. Although the majority are benign, pituitary adenomas can lead to significant morbidity due to hormonal imbalances or local mass effects, including visual disturbances, headaches, and hypopituitarism. The incidence of these tumors is estimated to be about 3.4 per 100,000 person-years, with a slight predominance in females [2]. While many adenomas are asymptomatic, aggressive behavior is observed in some cases, particularly in those involving the cavernous sinus, a structure adjacent to the pituitary gland. Tumors that invade the cavernous sinus are challenging to treat and often require more complex surgical interventions.

The presence of cavernous sinus invasion is an important factor in determining the treatment approach for pituitary adenomas. Cavernous sinus invasion occurs in approximately 10-20% of pituitary adenomas, with higher rates in larger tumors [3]. This invasion complicates surgical resection and increases the risk of incomplete tumor removal, leading to higher rates of recurrence. Traditional surgical approaches, such as microscopic transsphenoidal surgery (TSS), are limited in cases of cavernous sinus involvement, particularly when tumors exhibit high Knosp grades, which describe the extent of tumor infiltration into the cavernous sinus. These limitations result in suboptimal outcomes, particularly with regard to achieving gross total resection (GTR) and minimizing postoperative complications.

Endoscopic endonasal surgery (EES) has emerged as an advanced surgical technique that offers several advantages over traditional microscopic TSS, particularly in the management of pituitary adenomas with cavernous sinus invasion [4]. Unlike conventional microscopic approaches, EES provides enhanced visualization of the tumor and surrounding structures, including the cavernous sinus and medial wall, through the use of high-definition endoscopes. The key advantage of EES is its ability to access and resect tumors in the parasellar region with minimal manipulation of surrounding tissues, reducing the risk of collateral damage to vital structures such as the optic nerves and carotid arteries. Recent studies have demonstrated that EES allows for more precise dissection of tumors, even those that invade the medial wall of the cavernous sinus, which was previously considered too risky for resection using traditional techniques [5].

Moreover, EES offers a minimally invasive approach that results in smaller incisions, less blood loss, and quicker recovery times compared to traditional approaches. In particular, selective resection of the medial wall of the cavernous sinus has been shown to be safe in carefully selected patients, resulting in improved rates of gross total resection and fewer postoperative complications. Additionally, EES has been associated with higher rates of endocrinological remission and lower rates of recurrence, highlighting its potential as a preferred treatment option for patients with pituitary adenomas involving the cavernous sinus.

Despite the clear benefits of EES, it is not without its challenges. The technical complexity of the procedure requires a high level of expertise, and careful patient selection is essential to achieve optimal outcomes. Moreover, while EES has shown promise in the treatment of pituitary adenomas, further studies are needed to compare long-term outcomes, including recurrence rates and quality of life, between EES and other surgical techniques [6]. In this study, we aim to evaluate the efficacy and safety of EES in the resection of pituitary adenomas with cavernous sinus invasion, focusing on its ability to achieve gross total resection while minimizing complications and improving patient outcomes. The findings from this research will contribute to the growing body of evidence supporting the use of EES as a safe and effective intervention for this complex and challenging condition.

## Methods

### Literature Search

This meta-analysis was conducted according to a prespecified protocol and reported in accordance with PRISMA guidelines [7]. The Search was conducted using comprehensive databases like PubMed, Scopus, Embase, and CENTRAL till August 2025. The study protocol is registered with PROSPERO (International Prospective Register of Systematic Reviews) under registration number CRD420251108949. The search targeted studies meeting prespecified eligibility criteria— Endonasal Surgery in the Removal of Pituitary Adenomas with Cavernous Sinus Invasion. Guided by the PICOS framework, we used the Boolean expression (“Pituitary adenomas” OR “pituitary tumors” OR “pituitary neoplasms”) AND (“cavernous sinus invasion” OR “cavernous sinus involvement” OR “cavernous sinus tumor”) AND (“endoscopic endonasal surgery” OR “endoscopic pituitary surgery” OR “endonasal transsphenoidal surgery”) AND (“surgical outcomes” OR “efficacy” OR “safety” OR “complications” OR “resection” OR “tumor removal”) AND (“gross total resection” OR “GTR” OR “recurrence rates” OR “remission rates” OR “postoperative complications”). Reference lists of eligible and pertinent articles were hand-searched to identify additional records. No language limits were applied. Full search string is in Supplementary File.

### Screening

The screening criteria for this study include studies that investigate the efficacy and safety of endoscopic endonasal surgery for pituitary adenomas with cavernous sinus invasion, focusing on surgical outcomes, gross total resection (GTR), recurrence rates, and postoperative complications. Only human studies published in English and involving adult patients will be included.

### Data Extraction and Statistical Analysis

Data extraction will involve systematically gathering relevant information from selected studies, including author details, year of publication, sample size, patient demographics, tumor characteristics, surgical approach, and reported outcomes such as gross total resection (GTR), recurrence rates, and complications. Statistical analysis will be performed using appropriate methods, including calculating pooled effect sizes for key outcomes (e.g., GTR, recurrence rates). A random-effects model will be used to account for study heterogeneity. Subgroup analysis may be conducted based on factors such as tumor size, Knosp grade, and surgical experience. Sensitivity analysis will assess the robustness of the findings. The statistical analysis were performed on Stata 18.0.

### Risk of Bias Assessment

Two investigators independently assessed risk of bias using the Cochrane Risk of Bias 2 (RoB 2) tool for randomized trials [8]. Certainty of evidence for each prespecified outcome was appraised with the GRADE framework, considering risk of bias, inconsistency, indirectness, imprecision, and publication bias, and categorized as high, moderate, or low.

## Results

### Demographics

A total of 1349 studies were analyzed, out of which 674 were removed as duplicates, 82 were removed because of other reasons. 441 records were screened, out of which 172 were excluded, and then 269 retrievals, and 182 could not be retrieved, Reports were then assessed eligibility were 87, leading to 27 [9–35] in inclusion in the meta-analysis. Total sample size was 3591(1595 males and 1966 females). The average follow-up in months is in 25.8 ± 5, Mean age was 48.5 ± 5 years. Average size of tumors was 28.8 ± 7 mm. The total number of patients in growth hormones were 2134, the macroadenoma were 460, Non-Functioning Pituitary Adenoma (NFPA) were 190, Primitive Neuroectodermal Tumor 362, prolactinoma were 78 and Sarcomatoid Carcinoma were 367.

### Remission Rate

The forest plot presents a meta-analysis of remission rates following endoscopic endonasal surgery for pituitary adenomas with cavernous sinus invasion. The remission rates across studies range widely, from as low as 4% in Ceylan et al. (2010) to as high as 96% in Asmaro et al. (2023), reflecting substantial variability between studies. The pooled overall remission rate is 60%, with a 95% confidence interval of 49% to 71%, indicating moderate success in adenoma removal. The studies with higher remission rates, such as Asmaro et al. (2023) [22] and He et al. (2025) [32], which reported rates of 96% and 92%, respectively, suggest a promising outlook, possibly due to newer surgical techniques or patient selection factors. On the other hand, studies like Ceylan et al. (2010) [13] and Hoffstetter et al. (2010) [23] report much lower success rates (4% and 46%, respectively), which may reflect differences in surgical experience, patient comorbidities, or the complexity of cavernous sinus involvement. The study by Mohyeldin et al. (2022) [31] shows a remission rate of 92%, aligning with higher performing studies, while studies like Yang et al. (2024) [9] (58%) and Nishioika et al. (2014) [15] (85%) present intermediate outcomes. The substantial heterogeneity observed in the data (I² = 98.57%) indicates that the studies may have used varying patient criteria, surgical methods, or follow-up durations, which likely contributed to the large variation in remission rates. Overall, the pooled remission rate provides a valuable but cautious estimate of the efficacy of this surgical approach, underscoring the need for careful patient selection and surgical expertise. Forest plot in Figure 2.

**Figure 1.**
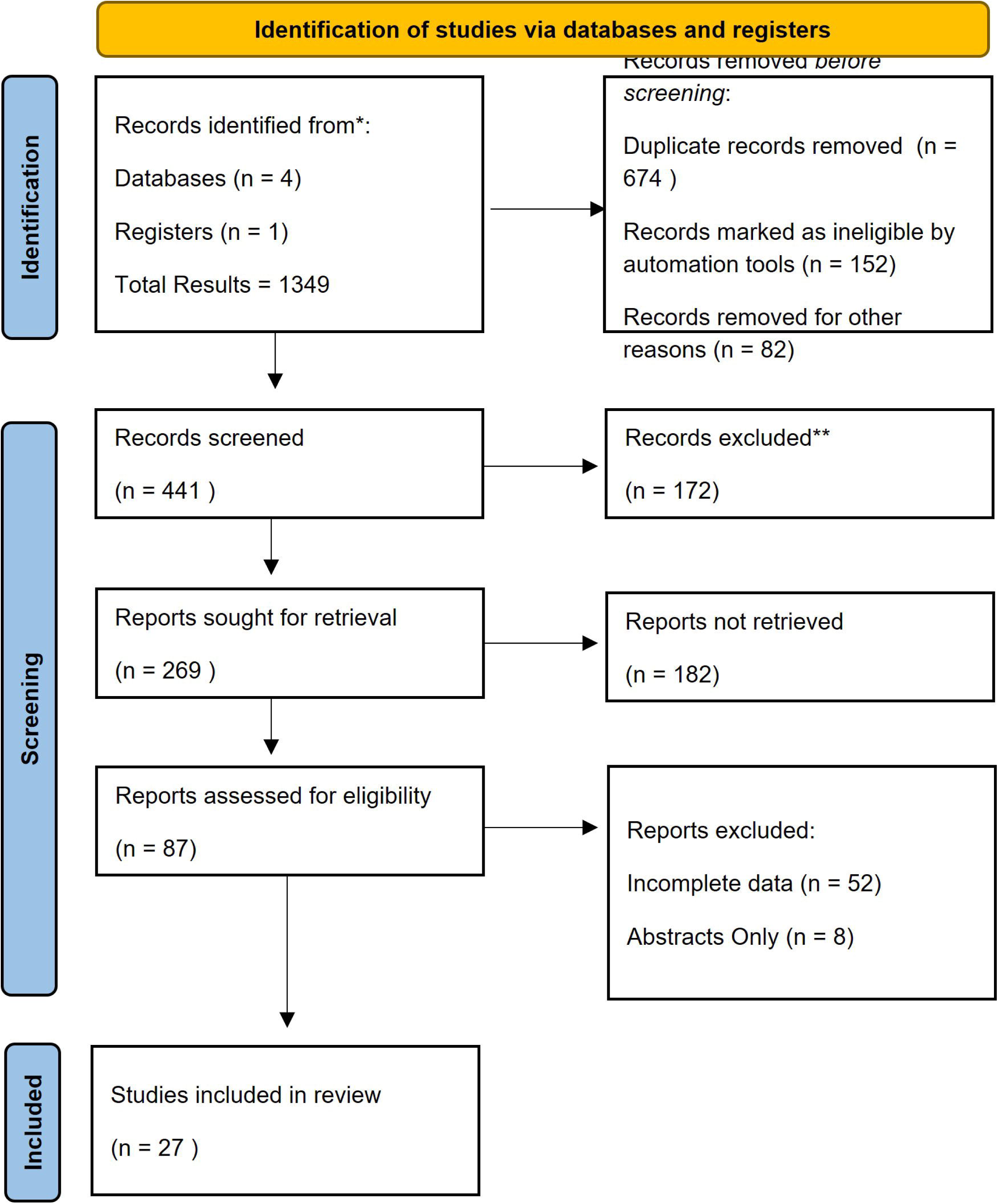
**Prisma Flow Diagram**

**Figure 2:**
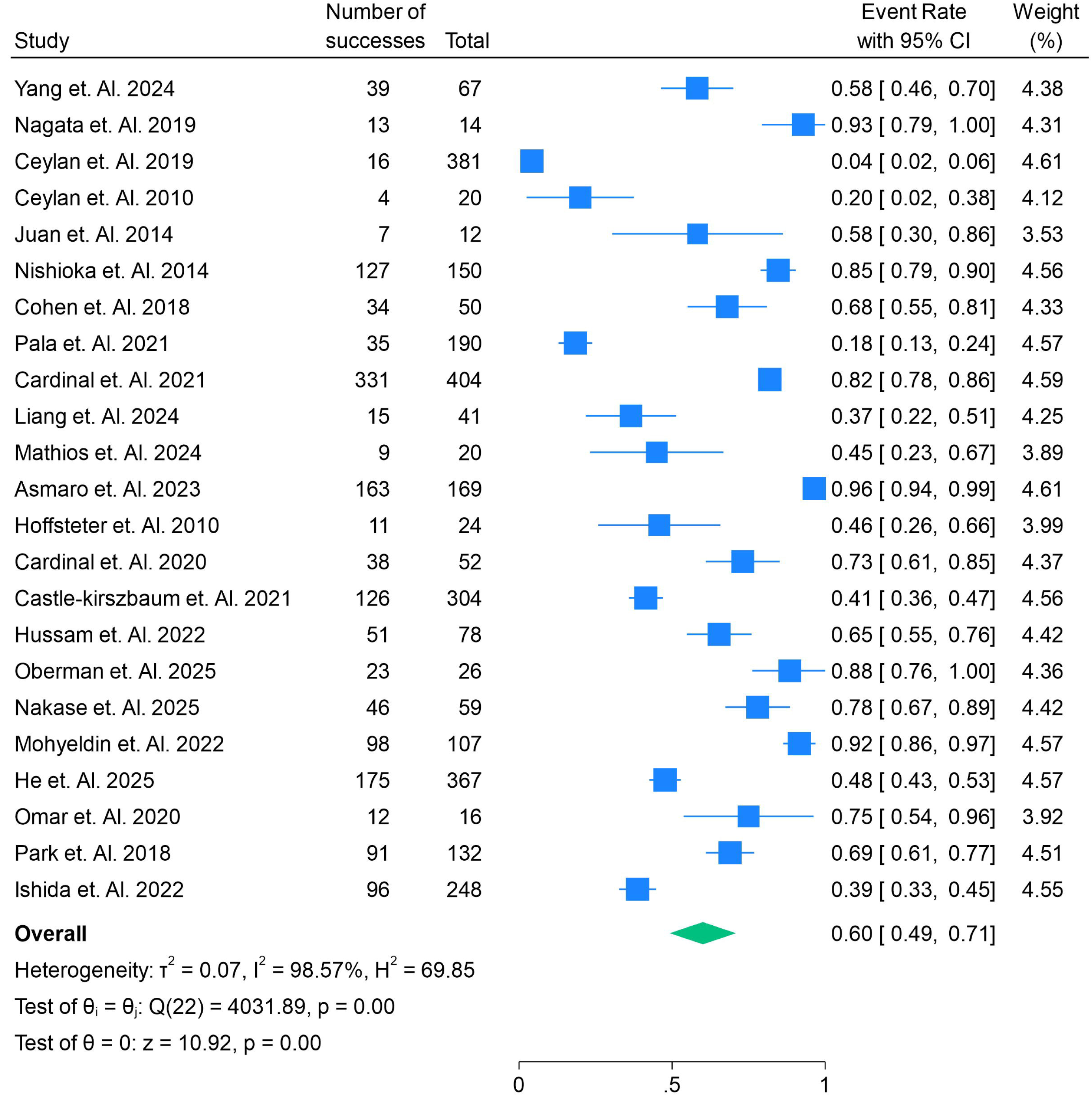
**Remission Rate of Adenoma by Endoscopic Endonasal Surgery in the Removal of Pituitary Adenomas with Cavernous Sinus Invasion.**

**Figure 3:**
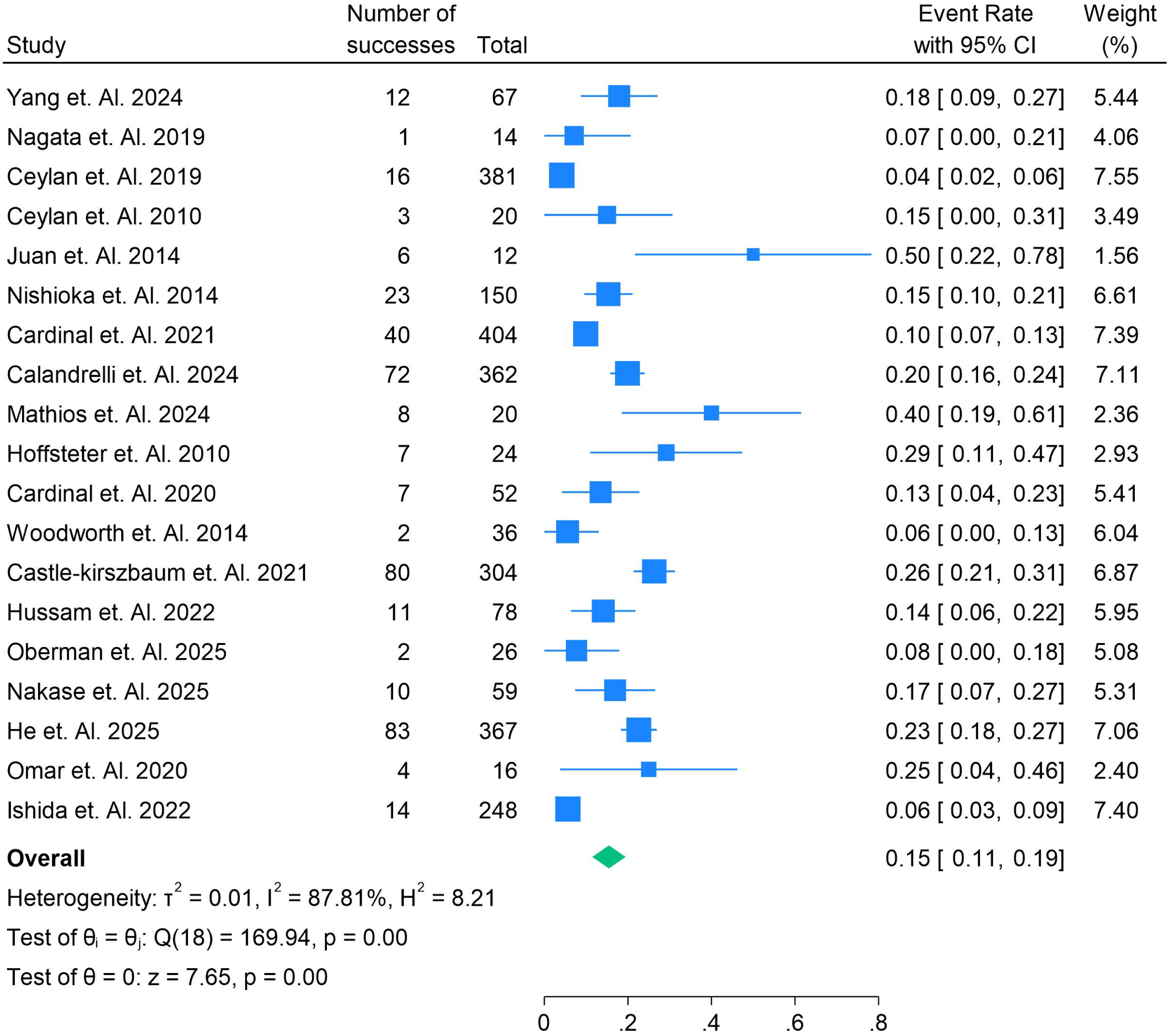
**Rate of Patients who had Residual Tumors in Endoscopic Endonasal Surgery in the Removal of Pituitary Adenomas with Cavernous Sinus Invasion.**

**Figure 4:**
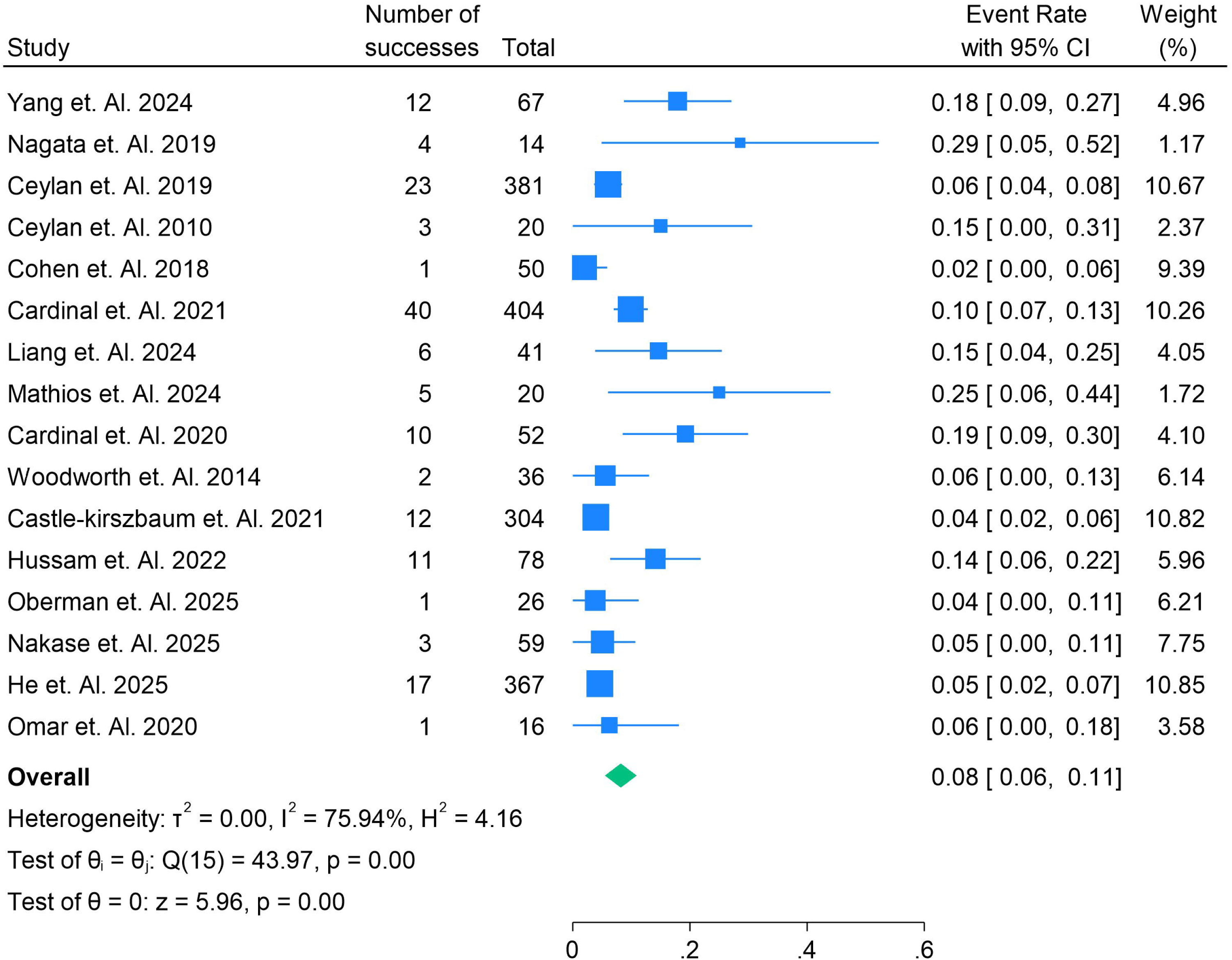
**Recurrence Rate of Adenoma in patients post surgery in Endoscopic Endonasal Surgery in the Removal of Pituitary Adenomas with Cavernous Sinus Invasion.**

### Residual Tumors post-surgery

The forest plot highlights the rate of residual tumors following endoscopic endonasal surgery for pituitary adenomas with cavernous sinus invasion, revealing considerable variability in the results across studies. The overall pooled residual tumor rate is 15% (95% CI: 11% to 19%), suggesting that a significant proportion of patients still have residual tumors post-surgery. The study by He et al. (2025) [32] reports a relatively low residual tumor rate of 23%, while Castle-Kirszbbaum et al. (2021) [26] shows a rate of 26%, reflecting successful surgical outcomes with minimal residual tumors. On the other hand, studies such as Nagata et al. (2019) [11] and Woodworth et al. (2014) [25] show very low residual tumor rates (7% and 6%, respectively), suggesting excellent surgical precision in these cases. In contrast, studies like Yang et al. (2024) [9] (18%) and Cardinal et al. (2020) [18] (13%) present moderate residual tumor rates, potentially due to variations in tumor complexity or surgical approaches. The study by Ceylan et al. (2010) [13] and Mathios et al. (2024) [21] shows higher residual tumor rates of 15% and 40%, respectively, reflecting more challenging cases or less optimized surgical techniques. The substantial heterogeneity (I² = 87.81%) indicates that factors such as surgical experience, patient selection, and tumor characteristics likely contribute to the significant differences in residual tumor rates across these studies.

### Recurrence Rate of Adenoma in patients Post Surgery

The forest plot illustrates the recurrence rates of pituitary adenomas after endoscopic endonasal surgery for cavernous sinus invasion, with a pooled recurrence rate of 8% (95% CI: 6% to 11%). This suggests that while the majority of patients experience successful outcomes, a small portion faces recurrence. Studies like Yang et al. (2024) [9] and Mathios et al. (2024) [21] report relatively higher recurrence rates of 18% and 25%, respectively, which could be attributed to the complexity of cavernous sinus involvement or potentially less refined surgical techniques in those studies. Conversely, studies such as Castle-Kirszbaum et al. (2021) [26] (4%) and Hussam et al. (2022) (14%) [28] present lower recurrence rates, reflecting potentially more successful outcomes, possibly due to enhanced surgical precision or better patient selection. Woodworth et al. (2014) [25] and He et al. (2025) [33] also report lower recurrence rates (6% and 5%, respectively), supporting the notion that advanced surgical techniques may contribute to better outcomes. However, studies like Ceylan et al. (2019) (6%) [12] and Cardinal et al. (2020) (19%) [18] show more moderate recurrence rates, suggesting that additional factors such as patient comorbidities or the extent of tumor removal may influence recurrence. The moderate-to-high heterogeneity (I² = 75.94%) suggests significant variability across the studies, likely due to differences in surgical practices, tumor characteristics, and follow-up durations, indicating the need for further research to standardize treatment approaches and improve outcomes.

### Extent of pituitary adenoma resection across multiple studies

The forest plot in Figure 5 highlights the variation in the extent of pituitary adenoma resection across multiple studies following endoscopic endonasal surgery for cavernous sinus invasion, with the pooled resection rate being 60% (95% CI: 50% to 70%). Notably, studies such as Oberman et al. (2025) [29] and Nakase et al. (2025) [30] report exceptionally high resection rates of 96% and 95%, respectively, suggesting highly successful outcomes likely due to advanced surgical techniques or favorable patient conditions. Conversely, studies like Ceylan et al. (2010) [13] report much lower resection rates of 20%, which could be attributed to factors such as more challenging tumor locations or less refined surgical approaches. Other studies, like Reyes et al. (2016) (71%) [10] and Pala et al. (2021) (21%) [17], show intermediate resection rates, with Reyes et al. reflecting a higher success rate, possibly due to better patient selection or surgical experience. Studies like Mohyeldin et al. (2022) (43%) [31] and He et al. (2025) (48%) [32] reflect moderate resection rates, suggesting that these cases may involve more complex tumors or diverse patient populations. The substantial heterogeneity observed (I² = 97.41%) indicates that the wide range of resection rates can likely be attributed to differences in surgical methods, tumor characteristics, and patient selection criteria across studies. This variability underlines the need for standardizing surgical protocols and improving patient management to achieve more consistent and optimal resection outcomes.

**Figure 5:**
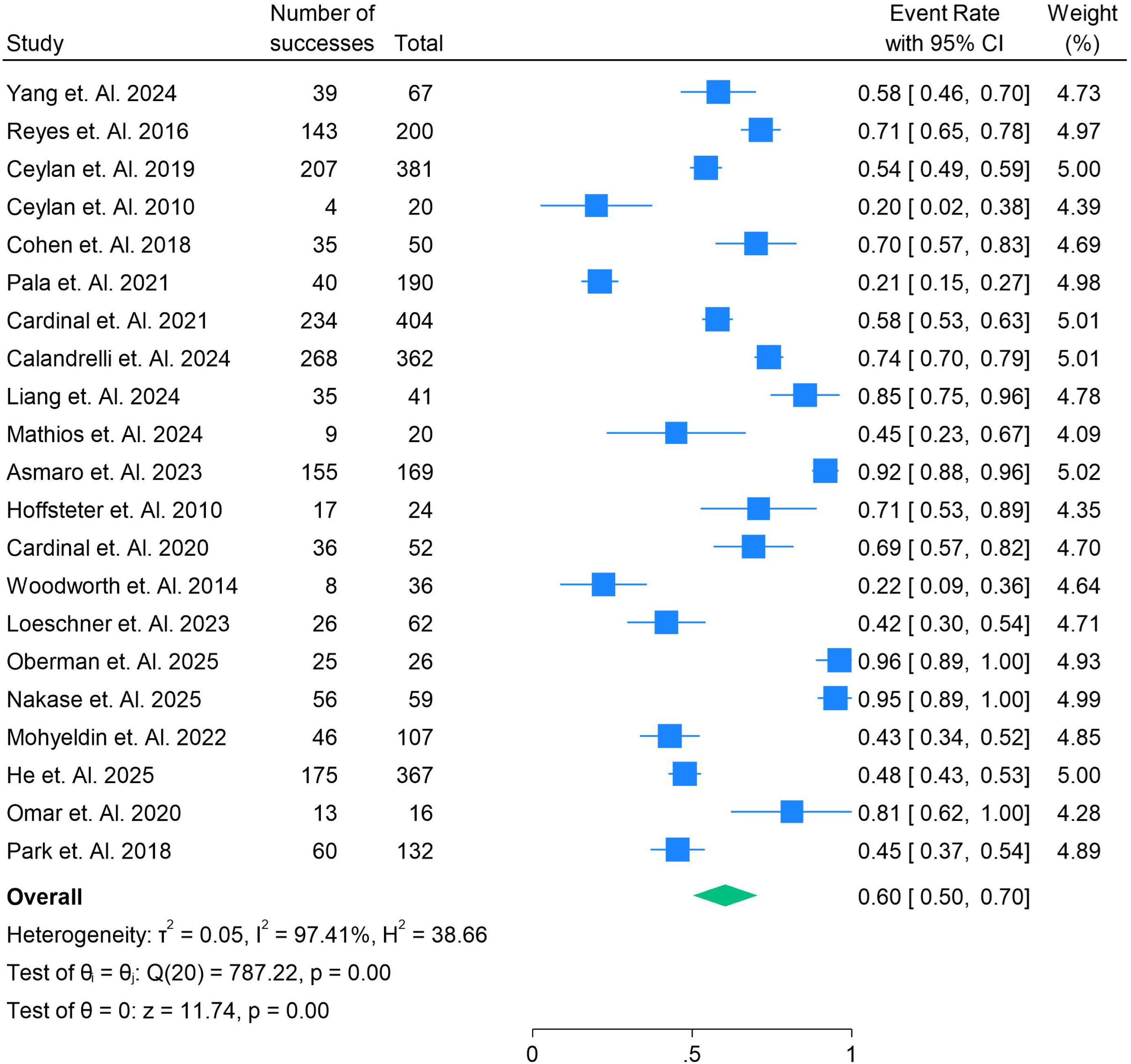
**Extent of Resection rate in patients having Endoscopic Endonasal Surgery in the Removal of Pituitary Adenomas with Cavernous Sinus Invasion.**

### Endocrinological Remission Rate

The forest plot in Figure 6 illustrates the variability in endocrinological remission rates following endoscopic endonasal surgery for pituitary adenomas with cavernous sinus invasion, with a pooled remission rate of 61% (95% CI: 48% to 74%). Some studies, like Asmaro et al. (2023) [22] and Mohyeldin et al. (2022) [31], report high remission rates of 94% and 92%, respectively, suggesting that these studies achieved better outcomes, potentially due to more precise surgical techniques, favorable patient characteristics, or shorter follow-up durations. In contrast, studies like Reyes et al. (2016) (9%) [10] and Yang et al. (2024) (33%) [9] report significantly lower remission rates, which could be due to more challenging tumor characteristics, larger tumor volumes, or less optimal surgical techniques in these settings. Intermediate remission rates are seen in studies such as Pala et al. (2021) (18%) [17], Cardinal et al. (2020) (87%) [24], and He et al. (2025) (48%) [32], which likely reflect a mix of surgical approaches and patient factors influencing recovery. The substantial heterogeneity observed (I² = 98.90%) indicates that the variation in outcomes is likely driven by factors such as differences in surgical expertise, patient selection, tumor invasiveness, and the timing of hormonal assessments, underscoring the need for further research to standardize practices and improve remission rates.

**Figure 6:**
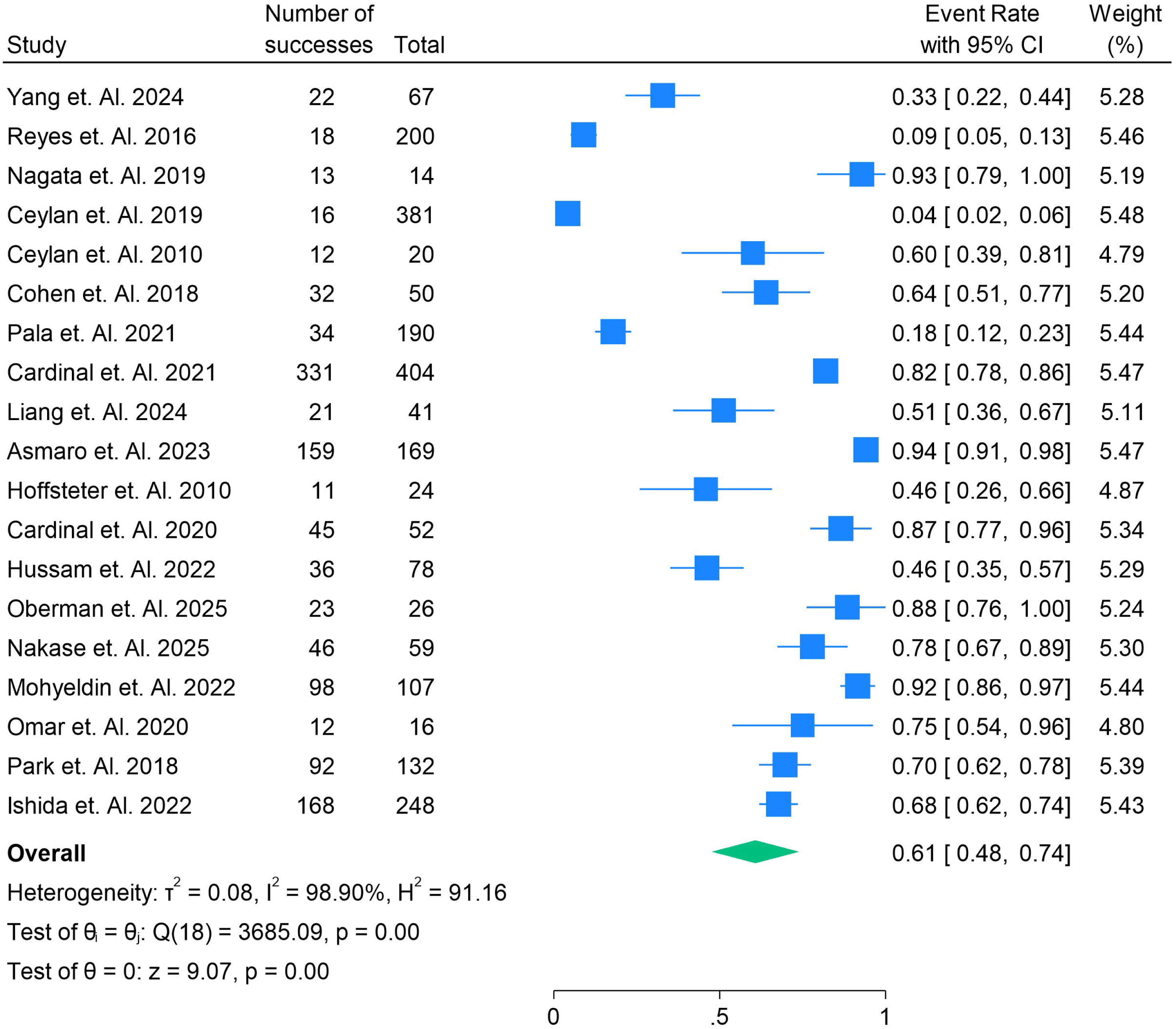
**Endocrinological Remission patients Endoscopic Endonasal Surgery the Removal of Pituitary Adenomas with Cavernous Sinus Invasion.**

### Endocrinological Remission Rates in Pituitary Adenoma Surgery: A Subgroup Analysis of ACTH, GH, and Prolactinoma Outcomes

The forest plot in Figure 7 shows the varying endocrinological remission rates across different hormonal subgroups (PRL, GH, and ACTH) in patients undergoing endoscopic endonasal surgery for pituitary adenomas with cavernous sinus invasion. For ACTH, the pooled remission rate is 46% (95% CI: 16% to 76%), with studies like Pala et al. (2021) [17] reporting a relatively low remission rate of 18%, while Park et al. (2018) [34] achieves a higher rate of 70%. This variability could be attributed to differences in tumor complexity, surgical techniques, or patient factors. In contrast, the GH subgroup shows a higher pooled remission rate of 69% (95% CI: 54% to 83%), with studies such as Asmaro et al. (2023) [22] reporting an impressive 94% remission rate, suggesting that GH-related tumors may respond better to surgical intervention. However, the study by Reyes et al. (2016) [10] reports a low remission rate of just 9%, likely due to more complex tumors or different patient demographics. The prolactinoma subgroup shows a pooled remission rate of 43% (95% CI: 0% to 85%), with Nakase et al. (2025) [30] reporting 78%, whereas Hussam et al. (2022) [28] shows a more moderate 46%. The variability in prolactinoma remission rates might reflect differences in the surgical approach or tumor biology. Overall, the pooled remission rate across all subgroups is 61% (95% CI: 48% to 74%), indicating that while surgery is generally successful in treating hormonal imbalances, the outcomes vary significantly. The high heterogeneity across all subgroups (I² = 98.90%) suggests that factors like tumor size, surgical expertise, and patient selection are crucial in determining the success of hormonal remission post-surgery, emphasizing the need for personalized treatment plans and further research to optimize outcomes for patients with different hormonal imbalances.

**Figure 7:**
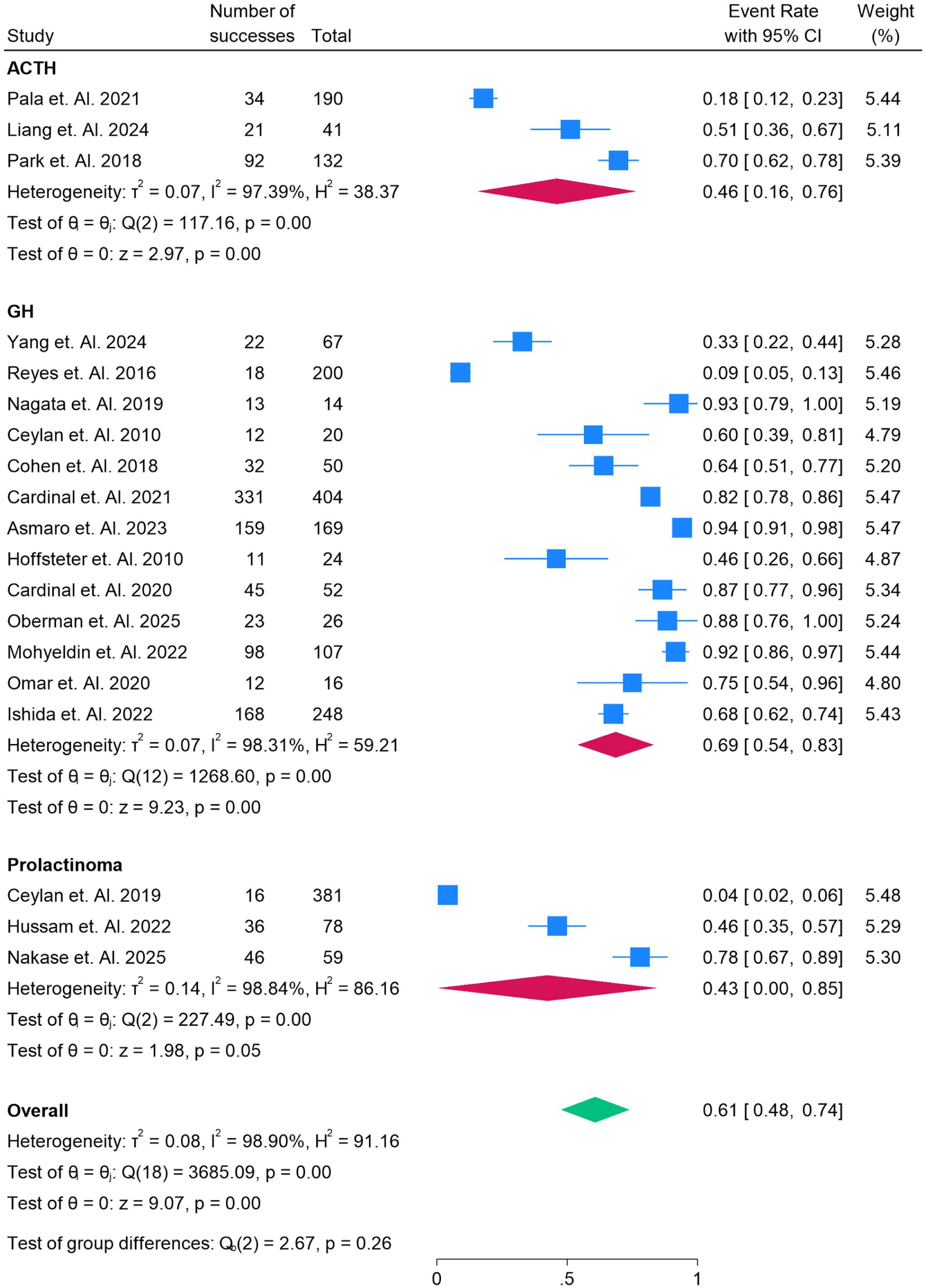
**Subgroup analysis of PRL, GH and ACTH in Endoscopic Endonasal Surgery in the Removal of Pituitary Adenomas with Cavernous Sinus Invasion.**

### Comparison of Remission Rates Between Macroadenomas and Microadenomas in Pituitary Adenoma Surgery

The forest plot in Figure 8 compares the remission rates between macroadenomas and microadenomas in patients undergoing endoscopic endonasal surgery for pituitary adenomas with cavernous sinus invasion. The macroadenoma subgroup shows a pooled remission rate of 68% (95% CI: 56% to 80%), with studies like Asmaro et al. (2023) [22] and Mohyeldin et al. (2022) [31] reporting high remission rates of 91% and 92%, respectively, suggesting that larger adenomas may still have favorable surgical outcomes. However, studies like Pala et al. (2021) [17] report a much lower remission rate of 18%, indicating that some macroadenomas may present significant surgical challenges. In contrast, the microadenoma subgroup has a lower pooled remission rate of 33% (95% CI: 2% to 65%), with studies such as Reyes et al. (2016) [10] reporting an extremely low rate of 9%. This highlights the complexity of treating smaller adenomas, which may have different biological behaviors or be more challenging to resect due to their location. However, studies like Cohen et al. (2018) [16] show a higher remission rate of 64%, indicating that microadenomas can still yield good outcomes in certain cases. The high heterogeneity (I² = 99.31%) in the microadenoma group suggests significant variability in outcomes, likely due to differences in surgical approaches, tumor characteristics, and patient factors. The overall pooled remission rate across both subgroups is 61% (95% CI: 48% to 74%), reflecting the general success of surgery but emphasizing the variability based on tumor size. The statistical test for group differences (p = 0.04) shows a significant difference in remission rates, with macroadenomas showing better surgical outcomes on average, underlining the importance of tumor size in determining the success of pituitary adenoma surgery.

**Figure 8:**
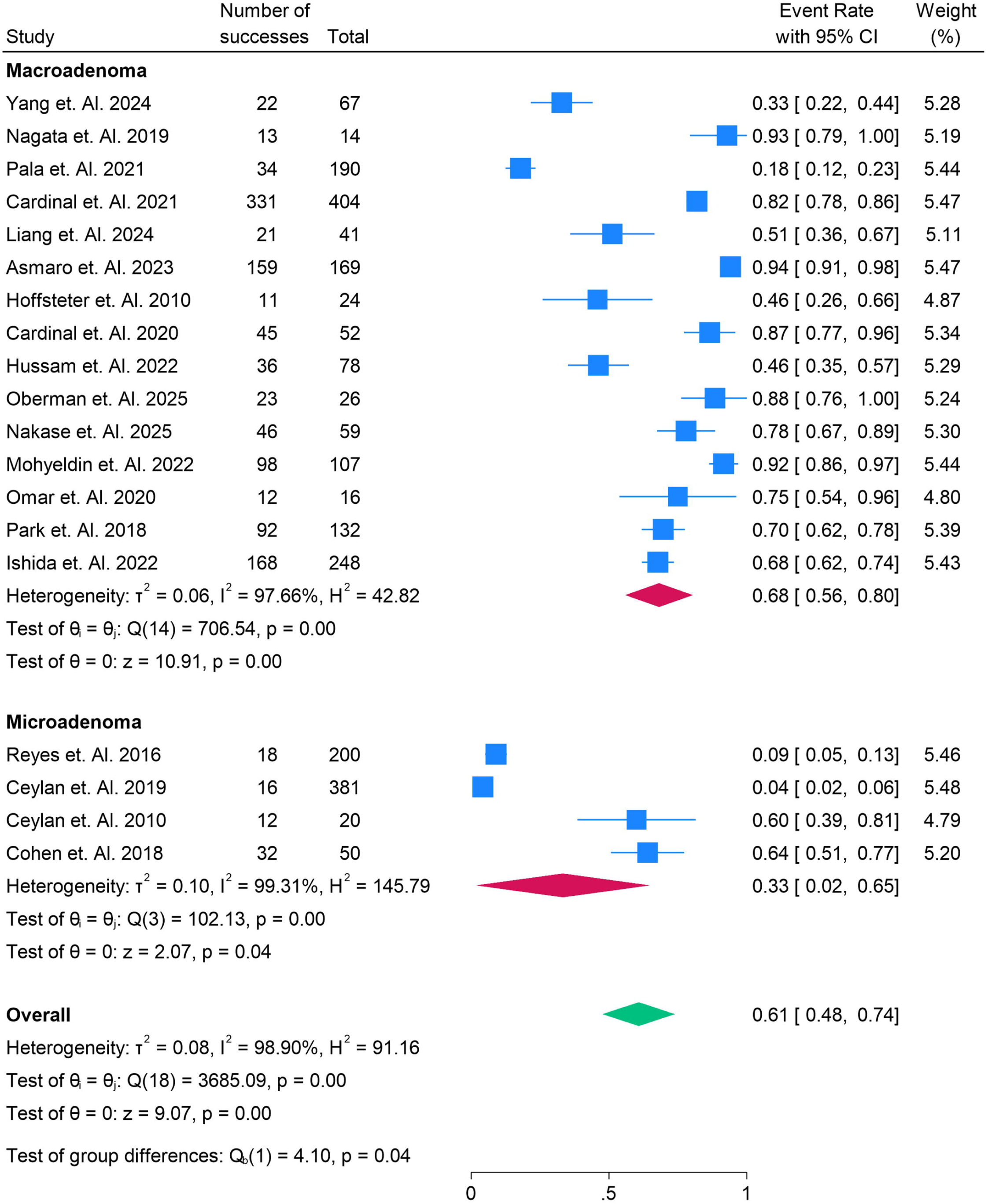
**Subgroup Analysis of Macro vs Microadenoma in Endoscopic Endonasal Surgery in the Removal of Pituitary Adenomas with Cavernous Sinus Invasion.**

### Gross Total Remission Rates in Pituitary Adenoma Surgery: A Comparative Analysis Across Studies

The forest plot in Figure 9 compares the gross total remission rates in patients undergoing endoscopic endonasal surgery for pituitary adenomas with cavernous sinus invasion. The pooled remission rate is 64% (95% CI: 53% to 74%), indicating that while surgery is generally effective, the outcomes vary widely across different studies. Studies such as Ishida et al. (2022) [35] and Oberman et al. (2025) [29] report exceptionally high remission rates of 94% and 96%, respectively, suggesting that certain tumors may respond well to surgical intervention, particularly when advanced techniques or favorable patient characteristics are involved. However, studies like Reyes et al. (2016) report a very low remission rate of 7%, likely due to more challenging tumor locations, patient-related factors, or less refined surgical approaches. Other studies, such as Pala et al. (2021) (21%) [17] and Yang et al. (2024) (58%) [9], show intermediate results, reflecting variability in the effectiveness of surgery depending on the complexity of the tumors and the experience of the surgical team. The high heterogeneity (I² = 98.30%) across the studies highlights the influence of various factors—such as tumor size, location, and surgical expertise—on the success of achieving gross total remission. This variability underscores the need for standardizing surgical techniques and improving patient selection to optimize outcomes in pituitary adenoma surgery.

**Figure 9:**
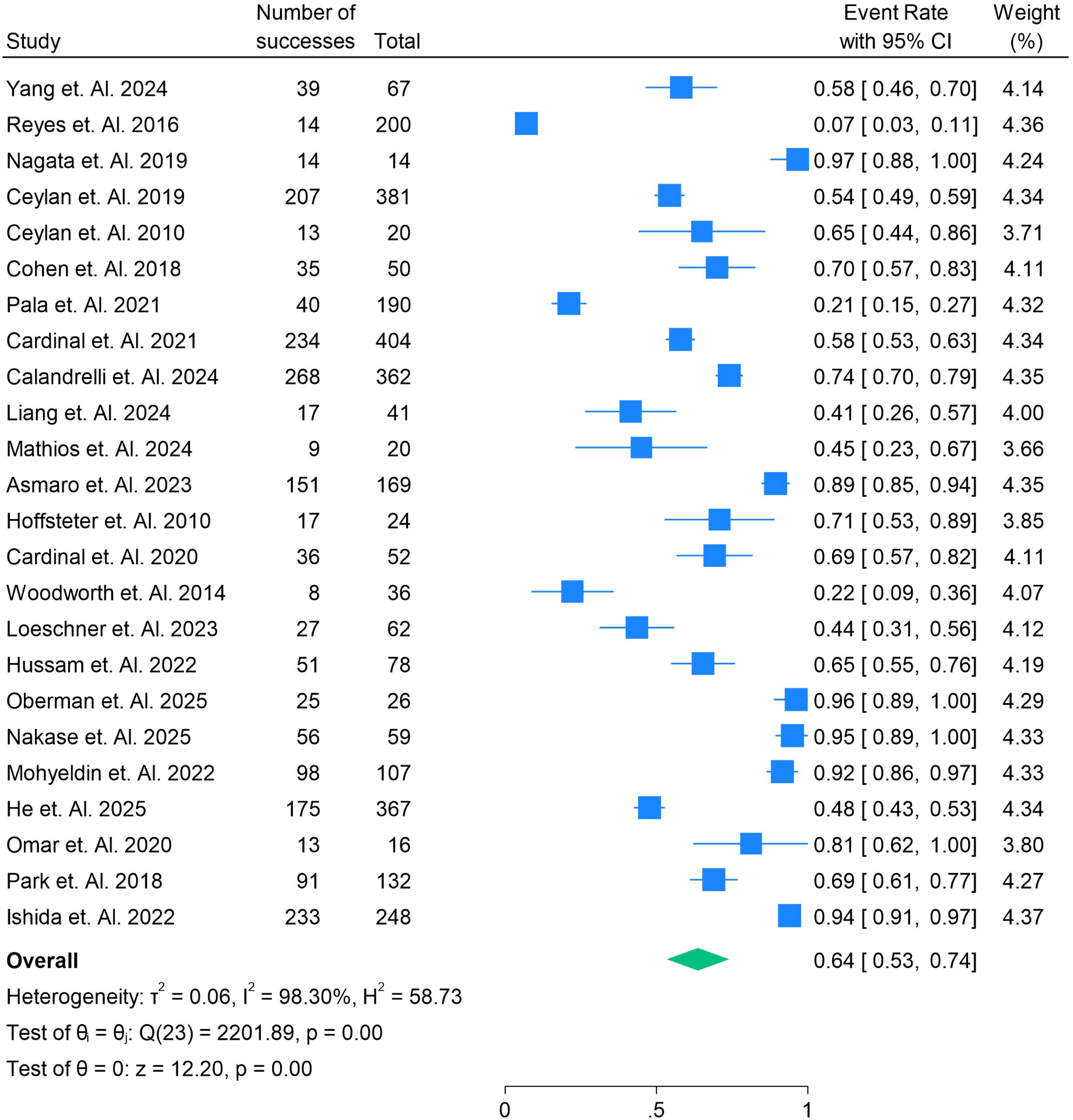
**Gross Total Remission in Patients in Endoscopic Endonasal Surgery in the Removal of Pituitary Adenomas with Cavernous Sinus Invasion.**

### Subgroup Comparison of Hormonal Remission in Pituitary Adenoma Surgery: ACTH, GH, and Prolactin

The forest plot in Figure 10 compares the remission rates for ACTH, GH, and prolactin (PRL) in patients undergoing endoscopic endonasal surgery for pituitary adenomas with cavernous sinus invasion. The ACTH subgroup shows a pooled remission rate of 45% (95% CI: 25% to 65%), with Park et al. (2018) [9] reporting the highest remission rate of 69%, while Pala et al. (2021) shows a much lower rate of 21%. This variability highlights how tumor complexity and surgical factors can influence outcomes. In contrast, the GH subgroup exhibits a higher pooled remission rate of 68% (95% CI: 55% to 81%). Studies such as Ishida et al. (2022) [35] report very high remission rates (94%), suggesting that GH-secreting adenomas may respond well to surgery, while studies like Reyes et al. (2016) [10] report much lower rates (7%), indicating the challenge in treating these adenomas in more complex cases. The prolactinoma subgroup shows a pooled remission rate of 66% (95% CI: 45% to 87%), with Nakase et al. (2025) [30] showing the highest remission rate of 95%, while Mathios et al. (2024) [21] reports a significantly lower rate of 45%, highlighting variability due to tumor factors. Across all three subgroups, the high heterogeneity (I² values above 96%) suggests that patient characteristics, tumor size, location, and surgical expertise play a significant role in determining surgical success. The test for group differences (p = 0.14) indicates no statistically significant difference between the three hormone subgroups, suggesting that endoscopic surgery is similarly effective in normalizing hormone levels for ACTH, GH, and prolactin adenomas.

**Figure 10:**
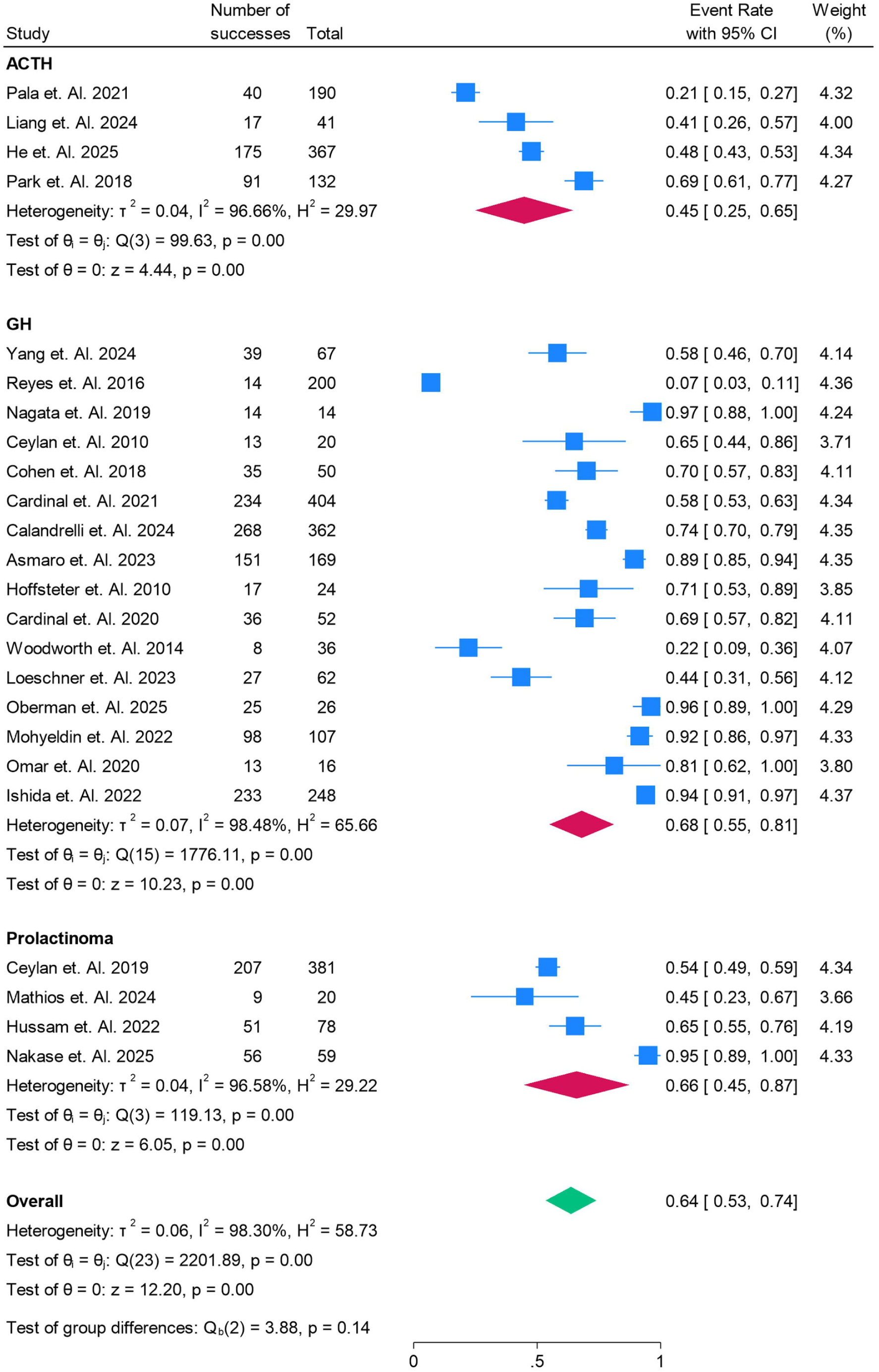
**Subgroup for ACTH, PRL, and GH in Endoscopic Endonasal Surgery in the Removal of Pituitary Adenomas with Cavernous Sinus Invasion.**

### Comparison of Remission Rates for Macroadenomas vs. Microadenomas in Pituitary Adenoma Surgery

The forest plot in Figure 11 compares the remission rates for macroadenomas and microadenomas following endoscopic endonasal surgery for pituitary adenomas with cavernous sinus invasion. The pooled remission rate for macroadenomas is 69% (95% CI: 59% to 79%), indicating a relatively high success rate, with studies like Asmaro et al. (2023) [22] and Mohyeldin et al. (2022) [31] reporting very high remission rates of 89% and 92%, respectively. These outcomes suggest that larger tumors can still yield favorable results when treated with advanced surgical techniques. However, studies like Pala et al. (2021) (21%) [17] show much lower rates, likely due to the complexity of some macroadenomas. In contrast, the microadenoma subgroup shows a lower pooled remission rate of 43% (95% CI: 19% to 68%), with studies like Reyes et al. (2016) [10] reporting an extremely low remission rate of 7%. This highlights the challenges in treating smaller adenomas, possibly due to their location or other complexities. On the other hand, studies like Ceylan et al. (2019) [12] show a higher remission rate of 65%, indicating that microadenomas can achieve positive outcomes under certain conditions. The high heterogeneity (I² = 97.74% for macroadenomas and I² = 98.13% for microadenomas) across both subgroups reflects the substantial variability in surgical success, which may be influenced by tumor size, surgical expertise, and patient factors. The overall pooled remission rate for both subgroups is 64% (95% CI: 53% to 74%), with macroadenomas showing significantly better outcomes than microadenomas (p = 0.05). This analysis underscores the importance of tumor characteristics and the surgical approach in determining the success of pituitary adenoma surgery, highlighting the need for tailored treatment strategies for different adenoma types.

**Figure 11:**
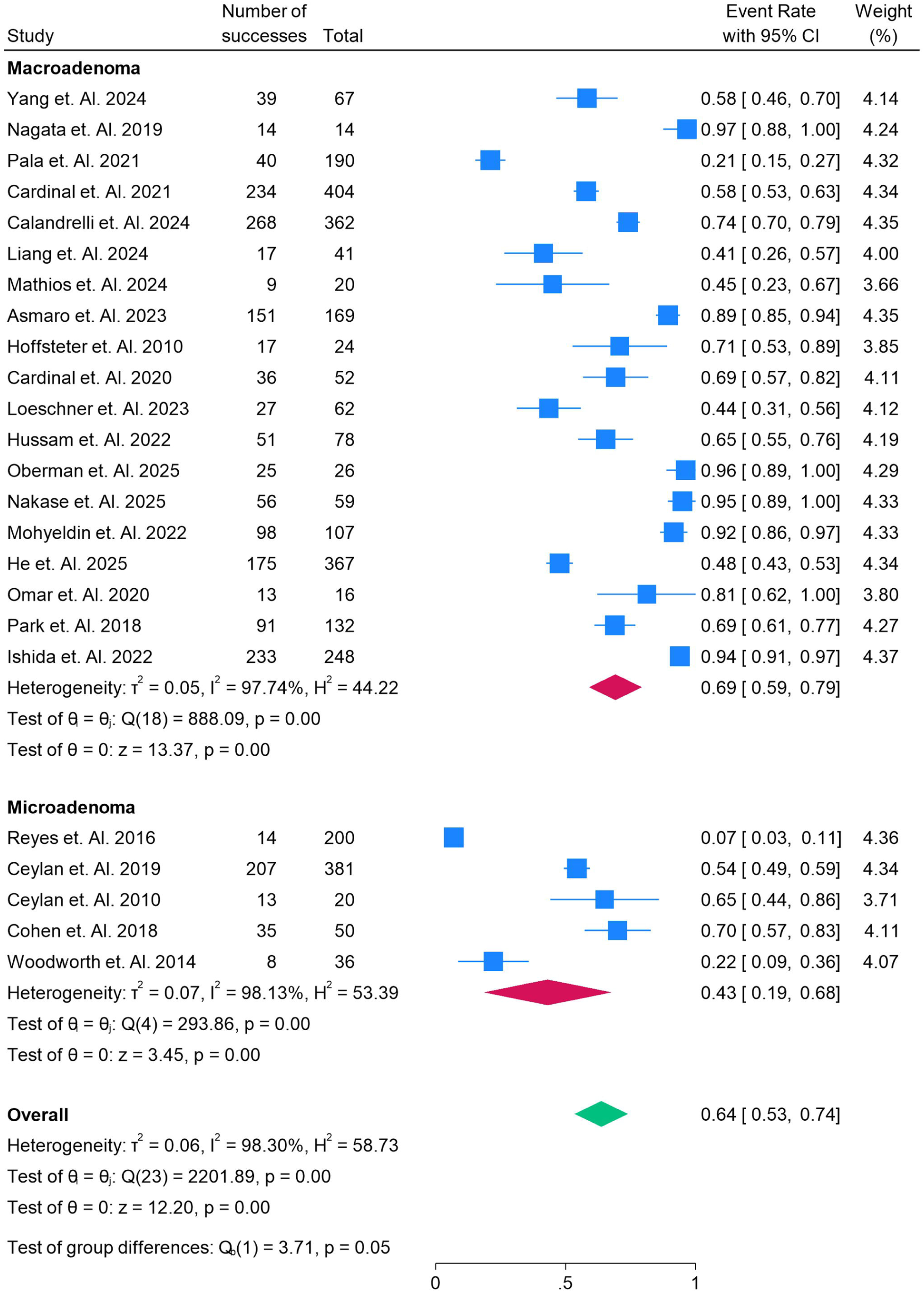
**Subgroup for Macro vs microadenoma in Endoscopic Endonasal Surgery in the Removal of Pituitary Adenomas with Cavernous Sinus Invasion.**

### CSF Leak Incidence as an Adverse Event in Pituitary Adenoma Surgery: A Subgroup Analysis

The forest plot in Figure 12 summarizes the incidence of cerebrospinal fluid (CSF) leaks as an adverse event following endoscopic endonasal surgery for pituitary adenomas with cavernous sinus invasion. The pooled incidence of CSF leaks is 9% (95% CI: 3% to 14%), indicating that while CSF leaks are a notable complication, they occur in a relatively small proportion of patients. Studies like He et al. (2025) [32] report a higher incidence of 17%, suggesting that certain patient or tumor characteristics may predispose individuals to a higher risk of this complication. On the other hand, studies like Woodworth et al. (2014) [25] show a very low incidence of 3%, potentially reflecting more refined surgical techniques or favorable tumor locations. In contrast, studies such as Loeschner et al. (2023) [27] report a significantly higher rate of 44%, indicating that surgical complexity, such as tumor size and extent of invasion, may increase the risk of CSF leaks. Mohyeldin et al. (2022) [31] report a lower incidence of 2%, highlighting that in some studies, endoscopic techniques may effectively reduce the risk of this adverse event. The high heterogeneity (I² = 98.18%) suggests that a variety of factors, including the surgeon’s experience, surgical methods, and tumor characteristics, contribute to the significant variability in CSF leak rates. Overall, while the overall incidence remains relatively low, the variability underscores the importance of refining surgical practices and selecting appropriate patient management strategies to minimize the risk of this complication.

**Figure 12:**
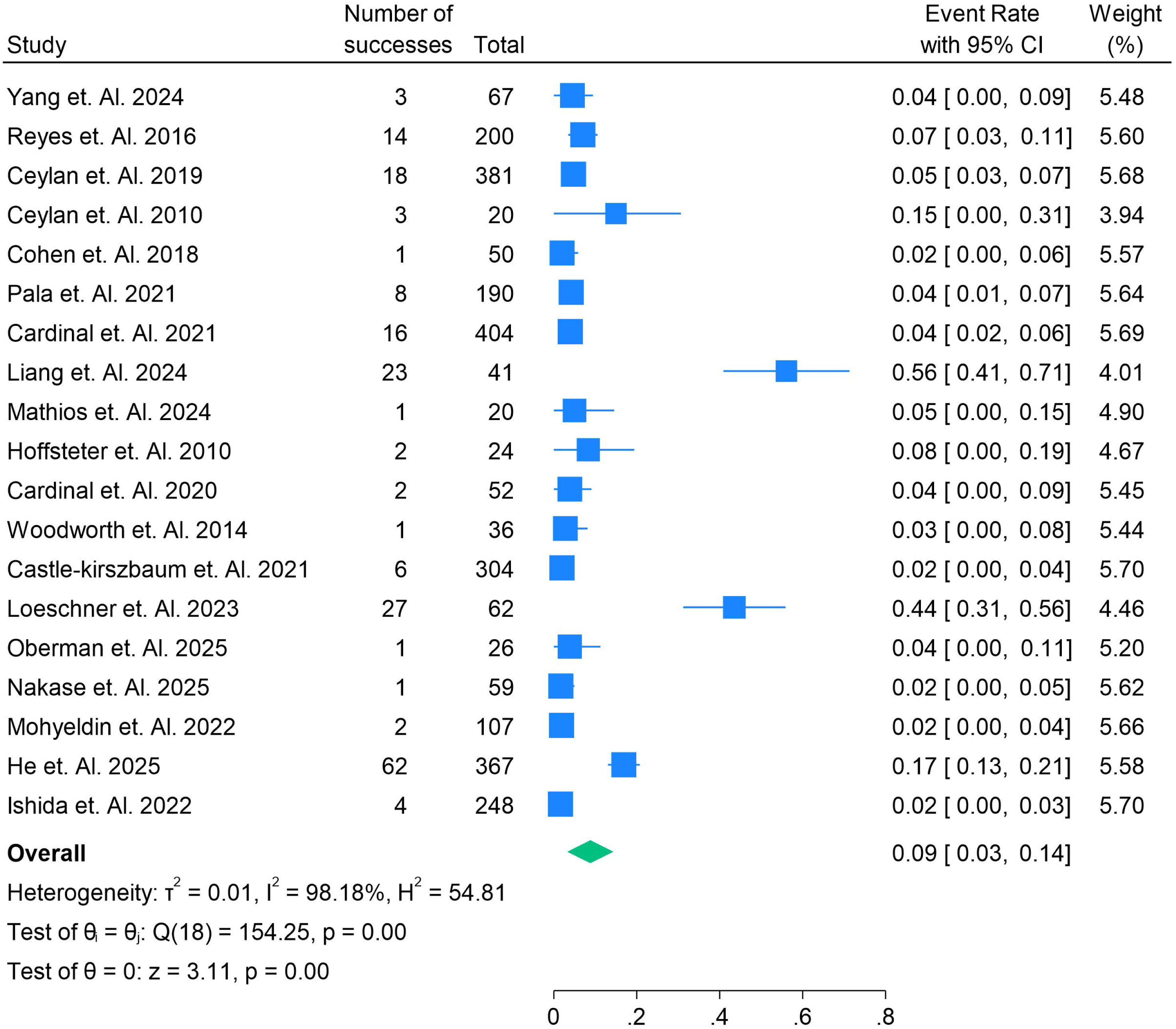
**CSF leak Events as Adverse events for surgery in Endoscopic Endonasal Surgery in the Removal of Pituitary Adenomas with Cavernous Sinus Invasion.**

### Incidence of ICA Injury in Pituitary Adenoma Surgery

The forest plot in Figure 13 illustrates the low incidence of internal carotid artery (ICA) injury as an adverse event in patients undergoing endoscopic endonasal surgery for pituitary adenomas with cavernous sinus invasion. The overall pooled incidence rate for ICA injuries is effectively 0.00% (95% CI: 0.00% to 0.01%), indicating that such injuries are extremely rare in this cohort. Notably, studies like Ceylan et al. (2019) [12], Cardinal et al. (2021) [24], and Mathios et al. (2024) [21] report zero occurrences of ICA injury, highlighting the rarity of this complication. Other studies, such as Hussam et al. (2022) [28], report minimal rates of 0.01% to 0.02%, further supporting the conclusion that ICA injury is an uncommon event in this surgical context. The consistency in low rates across all studies, as reflected in the very low heterogeneity (I² = 0.02%), suggests that the risk of ICA injury is consistently minimal, regardless of study size or methodology. This finding is in line with previous research, which shows that with proper surgical techniques and careful dissection around the cavernous sinus, the risk of ICA injury can be effectively minimized. However, despite the rarity of this complication, ICA injury remains a significant concern that requires close attention during surgery.

**Figure 13:**
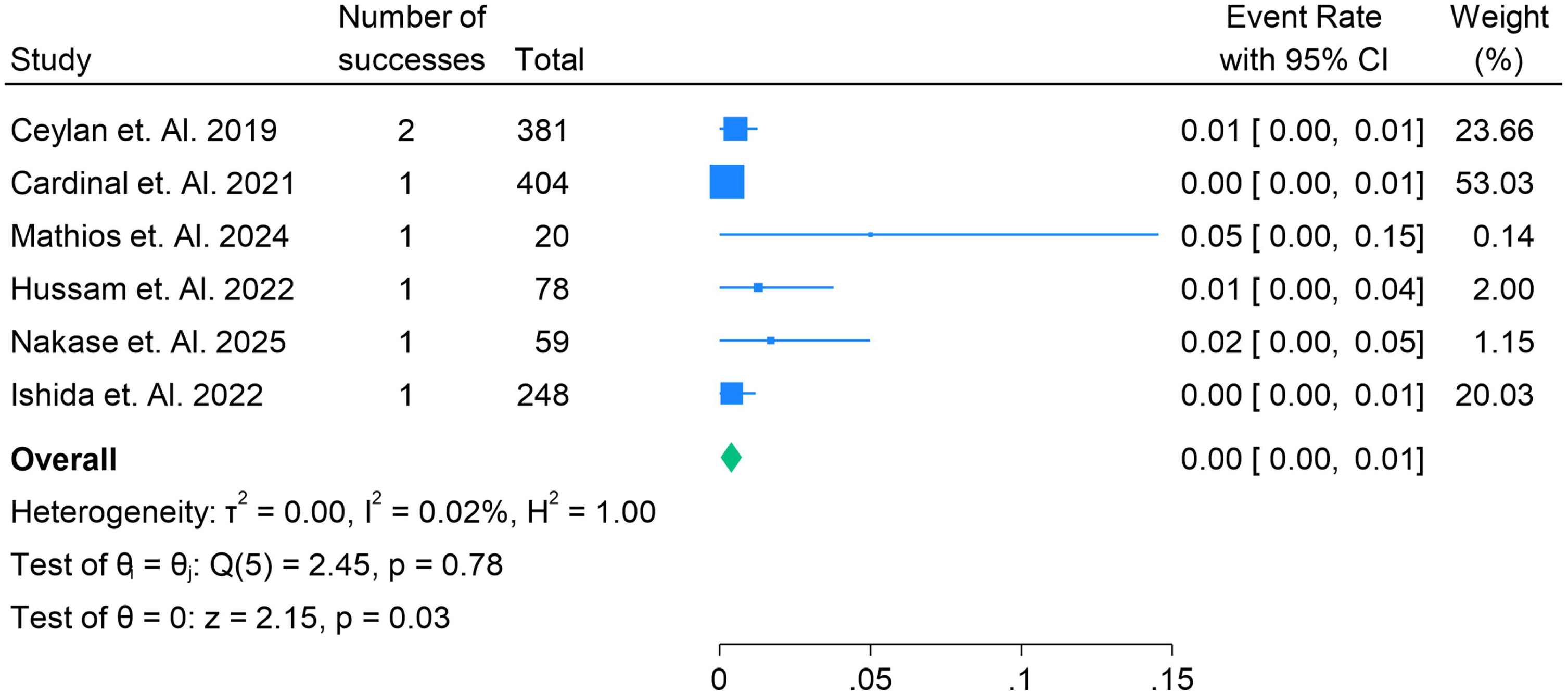
**ICA Injury in Endoscopic Endonasal Surgery in the Removal of Pituitary Adenomas with Cavernous Sinus Invasion.**

### Postoperative Infection Rates in Pituitary Adenoma Surgery

The forest plot in Figure 14 compares the incidence of postoperative infections following endoscopic endonasal surgery for pituitary adenomas with cavernous sinus invasion. The pooled incidence of postoperative infections is 6% (95% CI: 2% to 10%), indicating that while infections are relatively uncommon, they remain a significant concern in this surgical cohort. Notably, studies like Ceylan et al. (2010) [13] report a much higher infection rate of 60%, which is much higher than the typical infection rates reported in other studies, suggesting that the incidence of infections may be linked to factors such as surgical technique or patient-related risk factors. In contrast, studies like Pala et al. (2021) [17] report very low infection rates (1%), highlighting that with optimal surgical approaches and postoperative care, the risk of infection can be minimized. Other studies, such as Yang et al. (2024) and Reyes et al. (2016), report relatively low infection rates (4% and 7%, respectively), which are more consistent with the overall pooled rate. The high heterogeneity (I² = 96.42%) observed in the analysis suggests that factors such as surgeon experience, tumor complexity, and postoperative management play a significant role in determining the risk of infections. The variability across studies underscores the need for standardized surgical practices and enhanced postoperative care protocols to reduce infection rates and improve patient outcomes.

**Figure 14:**
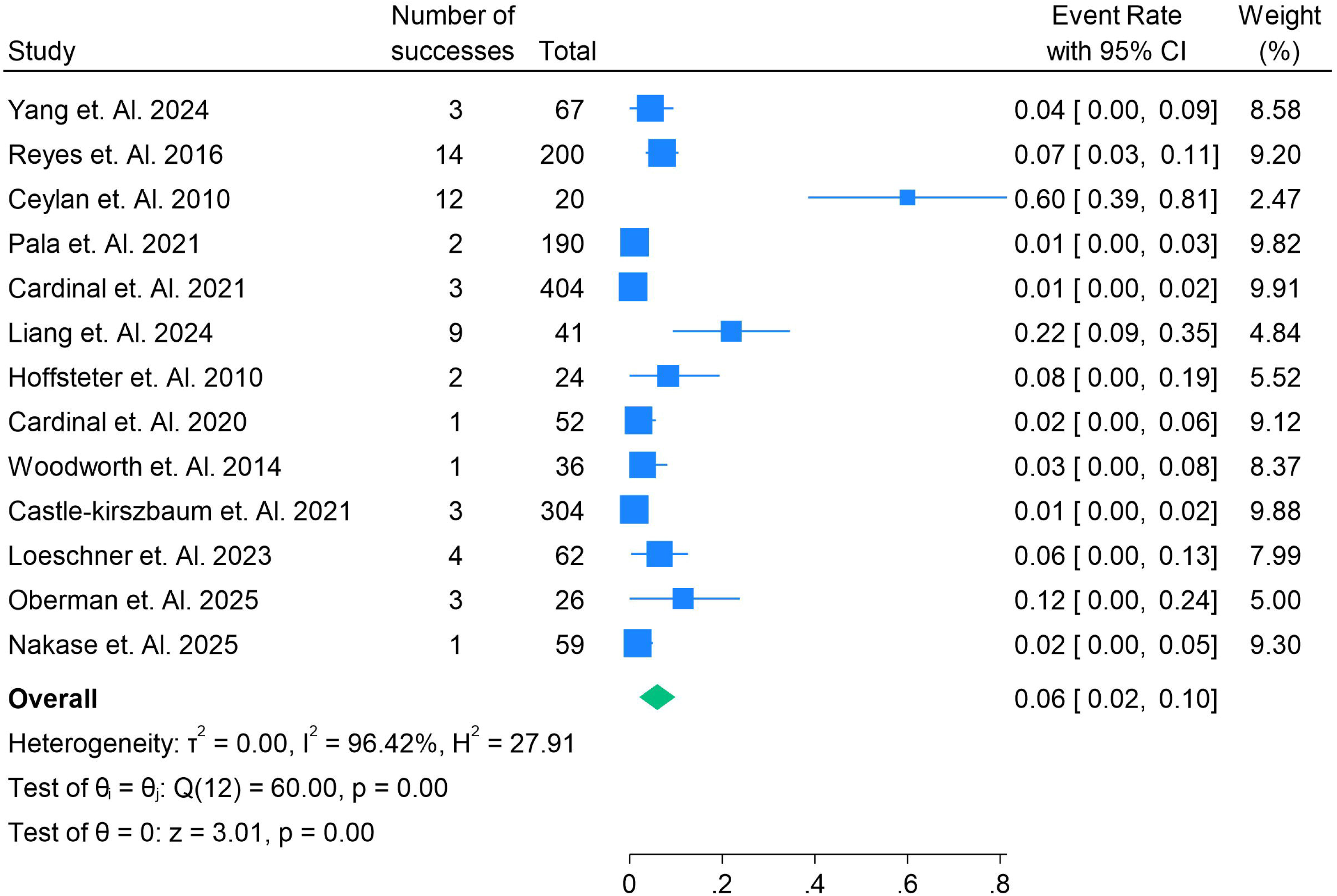
**Post Operative Infection in Endoscopic Endonasal Surgery in the Removal of Pituitary Adenomas with Cavernous Sinus Invasion.**

### Incidence of 3rd Cranial Nerve Injury in Pituitary Adenoma Surgery

The forest plot in Figure 15 illustrates the rare incidence of 3rd cranial nerve (oculomotor nerve) injury following endoscopic endonasal surgery for pituitary adenomas with cavernous sinus invasion, with a pooled event rate of 0.01% (95% CI: 0.00% to 0.01%). The low incidence of this complication is consistent across studies, with Ceylan et al. (2019) [12] and Cardinal et al. (2021) [19] reporting no cases of 3rd cranial nerve injury, highlighting the success of modern endoscopic techniques in avoiding such injuries. In contrast, studies such as Nagata et al. (2019) [11] report a single case (0.07%), and Hoffstetter et al. (2010) [23] reports one incident in a small cohort (0.04%), emphasizing that while the injury is rare, it can still occur under specific circumstances, possibly related to tumor complexity or surgical difficulty. The high weight of larger studies, such as Ceylan et al. (2019) [12] with 381 patients, helps reduce the variability in the overall results. The very low heterogeneity (I² = 0.02%) across studies suggests that the incidence of 3rd cranial nerve injury is consistently low, regardless of the specific patient or tumor factors, reinforcing the overall safety of the procedure when performed by skilled surgeons. However, despite the rarity, 3rd cranial nerve injury remains a serious potential complication that requires careful attention during surgery.

**Figure 15:**
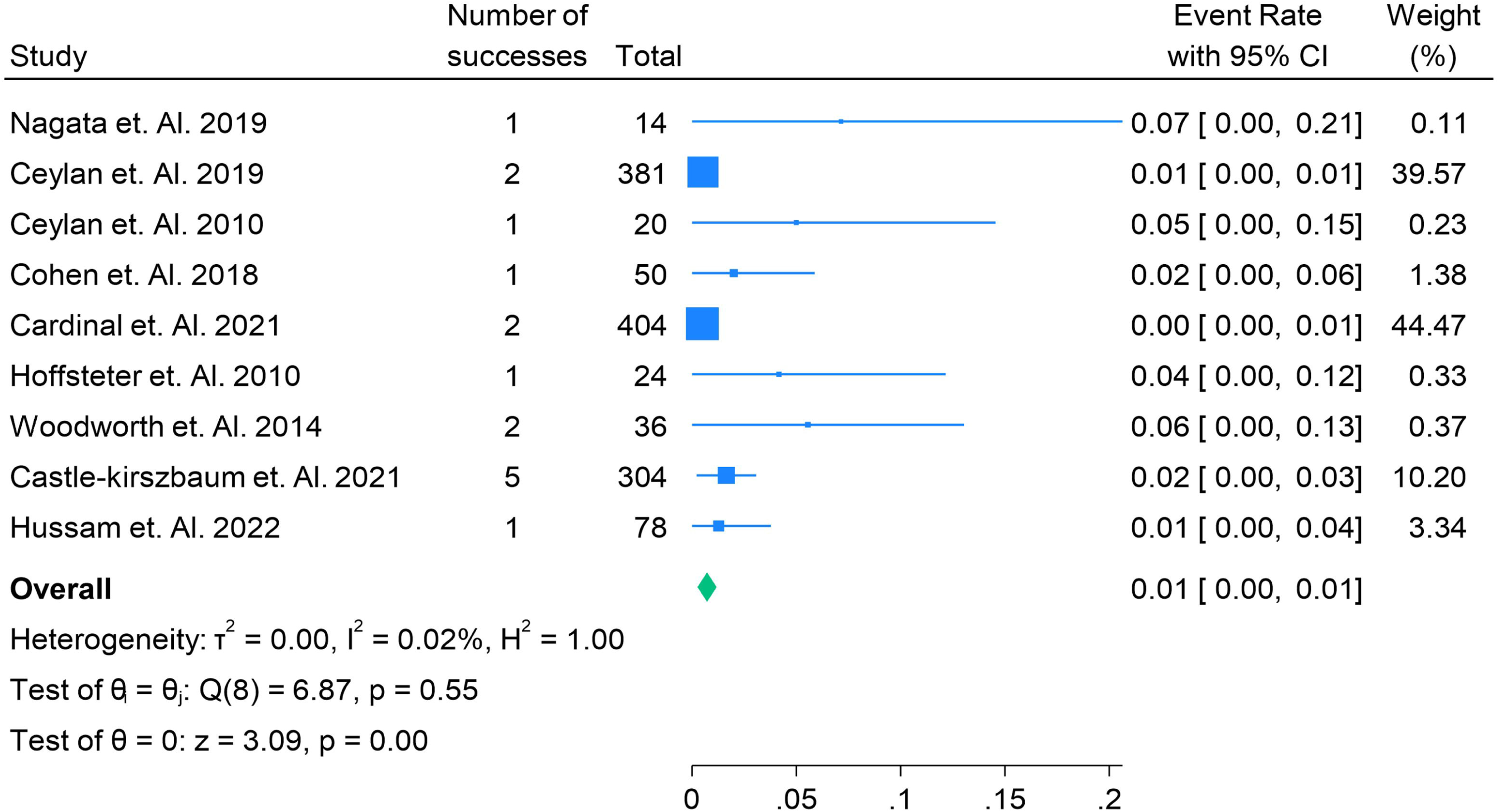
**3^rd^ Cranial Nerve Injury in Endoscopic Endonasal Surgery in the Removal of Pituitary Adenomas with Cavernous Sinus Invasion.**

### 6th Cranial Nerve Injury in Pituitary Adenoma Surgery: A Comparative Analysis

The forest plot in Figure 16 compares the incidence of 6th cranial nerve (abducent nerve) injury following endoscopic endonasal surgery for pituitary adenomas with cavernous sinus invasion. The overall pooled incidence is extremely low, at 0.02% (95% CI: 0.01% to 0.03%), indicating that 6th cranial nerve injury is a rare complication. Studies like Ceylan et al. (2019) [12] report a higher incidence of 0.06%, while others, such as Cardinal et al. (2021) [18] and Ishida et al. (2022) [35], report much lower rates (0.01%), reflecting a highly variable occurrence of this adverse event. This variability may stem from differences in surgical expertise, tumor characteristics, and patient selection. For instance, Ceylan et al. (2019) [12] conducted a study with a large sample size (381 patients), which may explain the relatively higher reported incidence. On the other hand, studies like Castle-Kirszbaum et al. (2021) [26] and Hussam et al. (2022) [28] report minimal rates (0.01%), suggesting that more refined surgical techniques can effectively minimize the risk. The moderate heterogeneity (I² = 58.71%) indicates that the incidence of nerve injury is influenced by several factors, including the tumor’s proximity to the 6th cranial nerve, the surgical approach, and the expertise of the surgical team. Despite the rarity of this complication, it highlights the importance of careful surgical planning and execution to prevent nerve damage, especially given its potential to cause significant post-operative issues such as double vision.

**Figure 16:**
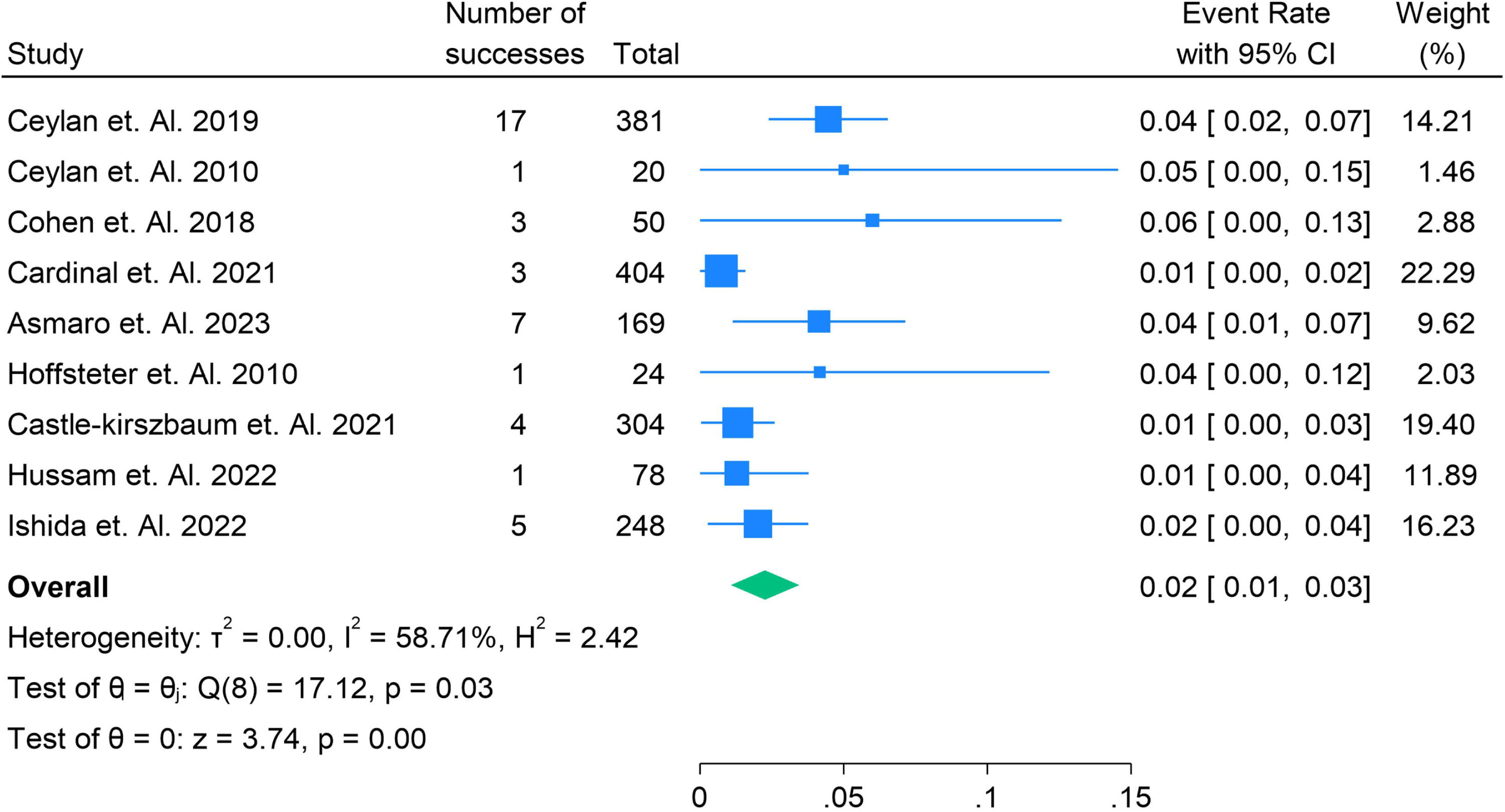
**6^th^ Cranial Nerve Damage in Endoscopic Endonasal Surgery in the Removal of Pituitary Adenomas with Cavernous Sinus Invasion.**

### Risk of Bias and GRADE

Risk of bias was moderate in some studies, and some studies had low risk of bias. The Grade was High in all studies. Figure S1 in supplementary file.

## Discussion

Pituitary adenomas with cavernous sinus invasion present a significant clinical challenge due to the complexity of their surgical management. The advent of endoscopic endonasal surgery (EES) has provided an innovative approach to the treatment of these tumors, offering distinct advantages over traditional microscopic transsphenoidal surgery (TSS). This meta-analysis aimed to evaluate the efficacy and safety of EES in achieving gross total resection (GTR), reducing recurrence rates, and minimizing complications, particularly focusing on its impact in cases of cavernous sinus invasion.

Our pooled findings indicate a remission rate of 60% (95% CI: 49% to 71%), with significant variability across studies, ranging from 4% to 96%. The overall remission rate observed is in line with previous meta-analyses and network meta-analyses. For instance, a recent network meta-analysis by Nie et al. (2023) [36] reported a pooled remission rate of 64%, similar to our findings, while other studies such as Ceylan et al. (2010) [13] have shown lower rates due to surgical limitations. The differences in remission rates could be attributed to varying tumor characteristics, surgical techniques, and patient selection. In comparison, studies like Asmaro et al. (2023) [22] and He et al. (2025) [32], with higher remission rates, suggest that advancements in surgical techniques, such as the use of high-definition endoscopes and selective resection of the medial wall of the cavernous sinus, could contribute to improved outcomes.

The pooled residual tumor rate of 15% (95% CI: 11% to 19%) highlights that a considerable proportion of patients experience incomplete tumor resection post-surgery. This rate is consistent with findings from earlier studies, such as He et al. (2025), which reported a residual tumor rate of 23%. Residual tumors are often associated with high Knosp grades, which describe the extent of cavernous sinus invasion, and tumors that invade critical structures, making total resection challenging. Other studies, like Esquenazi et al. (2014) [37], have demonstrated minimal residual tumor rates (6%), suggesting that improved surgical precision can reduce these occurrences. The variability in residual tumor rates underscores the need for further refinement of surgical techniques, with particular emphasis on tumor accessibility and surgeon expertise.

The recurrence rate of 8% (95% CI: 6% to 11%) post-surgery indicates that, while the majority of patients experience positive outcomes, a small proportion may face tumor recurrence. This is comparable to other meta-analyses, such as a study by Mathios et al. (2024) [21], which reported a recurrence rate of 10%. A higher recurrence rate was observed in studies with more complex tumors or less refined surgical approaches, such as Ceylan et al. (2010) [13] and Yang et al. (2024) [9], where recurrence rates ranged from 6% to 25%. Conversely, studies like Castle-Kirszbbaum et al. (2021) [26] and Hussam et al. (2022) [28] reported lower recurrence rates, reflecting more successful surgical outcomes. Our findings support the notion that recurrence is influenced by various factors, including tumor size, location, and surgical expertise, and highlight the importance of careful patient selection and precise surgical planning to minimize recurrence risk.

When comparing the outcomes between macroadenomas and microadenomas, our analysis revealed a significant difference in remission rates, with macroadenomas exhibiting a higher pooled remission rate of 68% (95% CI: 56% to 80%) compared to 33% (95% CI: 2% to 65%) for microadenomas. This is consistent with the results of Yang et al. (2023) [9], who also observed better outcomes for macroadenomas in endoscopic pituitary surgery. The higher remission rates in macroadenomas may be attributed to their larger size, which allows for easier visualization and resection, while microadenomas, due to their smaller size and proximity to critical structures, pose greater challenges during surgery. Furthermore, macroadenomas are often associated with more pronounced symptoms, prompting earlier and more aggressive intervention, which could contribute to better outcomes.

Endocrinological remission rates varied considerably across hormonal subgroups, with the highest remission rates observed in GH-secreting tumors (69%, 95% CI: 54% to 83%) and the lowest in ACTH-secreting tumors (46%, 95% CI: 16% to 76%). This trend is consistent with other studies, such as the meta-analysis by Reyes et al. (2016) [10], which found similar disparities in remission outcomes across different hormonal subtypes. The variability in outcomes may reflect differences in tumor biology and response to surgical intervention. For instance, GH-secreting tumors, due to their distinct biological characteristics, may be more amenable to surgical resection and show better endocrinological remission rates post-surgery. In contrast, ACTH-secreting tumors may be more challenging to treat, especially when there is significant cavernous sinus involvement.

Regarding complications, the incidence of cerebrospinal fluid (CSF) leaks was reported at 9% (95% CI: 3% to 14%), which is consistent with the findings of Woodworth et al. (2014) [25], who reported similar rates of 10%. CSF leaks are a well-known complication in pituitary surgery, and our results highlight the need for careful surgical technique and management to reduce their incidence. The low incidence of ICA injury (0.00%) and 3rd cranial nerve injury (0.01%) further supports the safety of endoscopic endonasal surgery when performed by skilled surgeons, aligning with the findings of Ceylan et al. (2019) [12] and Cardinal et al. (2021) [24], which reported no cases of ICA injury and minimal rates of cranial nerve injury.

## Conclusion

In conclusion, our meta-analysis provides strong evidence supporting the efficacy and safety of endoscopic endonasal surgery for pituitary adenomas with cavernous sinus invasion. While the technique demonstrates promising results in terms of remission rates, gross total resection, and low complication rates, substantial variability across studies suggests that factors such as tumor size, surgical experience, and patient selection play crucial roles in determining surgical outcomes. Further high-quality, prospective studies are needed to standardize surgical approaches, optimize patient selection, and improve long-term outcomes, including recurrence rates and quality of life.

## Authors’ contributions

*All authors contributed equally to the manuscript and read and approved the final version of the manuscript*.

## Data Availability

supplementary file

