## supplementary file for "Endoscopic Endonasal Surgery for Pituitary Adenomas with Cavernous Sinus Invasion: A Comprehensive Meta-Analysis of Efficacy, Remission Rates, Surgical Outcomes, and Complications"

Supplementary Files

Search String

("Endoscopic endonasal surgery" OR "endonasal endoscopic surgery") AND ("pituitary adenoma" OR "pituitary tumor") AND ("cavernous sinus invasion" OR "cavernous sinus encroachment") AND ("efficacy" OR "safety" OR "outcomes" OR "complications") AND ("surgical removal" OR "surgical resection" OR "surgical intervention") AND ("pituitary adenoma resection" OR "endoscopic pituitary surgery")

Table S1. Demographics table

| **References** | **Author Year** | **Country** | **Total Population** | **Male** | **Female** | **Study Design** | **Follow Up in Months** | **Mean Age** | **Tumor Type** | **Size of Tumor in mm** |
| --- | --- | --- | --- | --- | --- | --- | --- | --- | --- | --- |
| 9 | Yang et. Al. 2024 | South Korea | 67 | 36 | 31 | Retrospective cohort | 53 | 51 | NFPA | 43 |
| 10 | Reyes et. Al. 2016 | Spain | 200 | 119 | 81 | Retrospective cohort | 51 | 47 | GH | 26 |
| 11 | Nagata et. Al. 2019 | Japan | 14 | 7 | 7 | Retrospective cohort | 11.8 | 57.9 | FPA | 13.8 |
| 12 | Ceylan et. Al. 2019 | Turkey | 381 | 199 | 182 | Retrospective cohort | 34 | 46.2 | Macroadenoma | 47 |
| 13 | Ceylan et. Al. 2010 | Turkey | 20 | 9 | 11 | Retrospective cohort | 45 | 65 | GH | 18 |
| 14 | Juan et. Al. 2014 | USA | 12 | 8 | 4 | Retrospective cohort | 6 | 19 | FPA | 19 |
| 15 | Nishioka et. Al. 2014 | Japan | 150 | 77 | 73 | Retrospective cohort | 22 | 47 | GH | 17.8 |
| 16 | Cohen et. Al. 2018 | USA | 50 | 25 | 25 | Retrospective cohort | 30 | 48 | FPA | 15.9 |
| 17 | Pala et. Al. 2021 | Germany | 190 | 106 | 84 | Retrospective cohort | 47 | 55 | NFPA | 30.8 |
| 18 | Cardinal et. Al. 2021 | USA | 404 | 160 | 244 | Retrospective cohort | 19 | 50.4 | FPA | 23 |
| 19 | Calandrelli et. Al. 2024 | Italy | 362 | 186 | 176 | Prospective Cohort | 9 | 53.4 | PitNETS | 35.8 |
| 20 | Liang et. Al. 2024 | China | 41 | 19 | 22 | Retrospective cohort | 12 | 46.9 | GH | 27 |
| 21 | Mathios et. Al. 2024 | USA | 20 | 8 | 12 | Retrospective cohort | 21 | 63 | Macroadenoma | 53 |
| 22 | Asmaro et. Al. 2023 | USA | 169 | 91 | 78 | Retrospective cohort | 3 | 52 | FPA | 52 |
| 23 | Hoffsteter et. Al. 2010 | USA | 24 | 13 | 11 | Prospective Cohort | 23 | 50.7 | GH | 15 |
| 24 | Cardinal et. Al. 2020 | USA | 52 | 16 | 36 | Retrospective cohort | 24 | 50.7 | FPA | 16 |
| 25 | Woodworth et. Al. 2014 | USA | 36 | 22 | 14 | Prospective Cohort | 13 | 47.3 | GH | 13 |
| 26 | Castle-kirszbaum et. Al. 2021 | Asutralia | 304 | 143 | 161 | Prospective Cohort | 12 | 53.8 | GH | 19 |
| 27 | Loeschner et. Al. 2023 | Germany | 62 | 46 | 16 | Retrospective cohort | 48 | 57.6 | FPA | 64 |
| 28 | Hussam et. Al. 2022 | USA | 78 | 46 | 32 | Retrospective cohort | 66.4 | 37 | Prolactinoma | 14 |
| 29 | Oberman et. Al. 2025 | USA | 26 | 13 | 13 | Prospective Cohort | 10 | 49.5 | GH | 18.44 |
| 30 | Nakase et. Al. 2025 | USA | 59 | 37 | 22 | Retrospective cohort | 19 | 36 | Macroadenoma | 36 |
| 31 | Mohyeldin et. Al. 2022 | USA | 107 | 45 | 62 | Prospective Cohort | 15.6 | 40.9 | FPA | 21 |
| 32 | He et. Al. 2025 | China | 367 | 44 | 323 | Retrospective cohort | 40.9 | 50.1 | SCA | 16.2 |
| 33 | Omar et. Al. 2020 | Canada | 16 | 3 | 13 | Cohort Study | 11 | 40.9 | GH | 28 |
| 34 | Park et. Al. 2018 | South Korea | 132 | 56 | 76 | Retrospective cohort | 40 | 42.2 | FPA | 33 |
| 35 | Ishida et. Al. 2022 | Japan | 248 | 61 | 187 | Retrospective cohort | 12 | 44.5 | FPA | 48 |

Figure S1. Risk of Bias with ROBINS-I

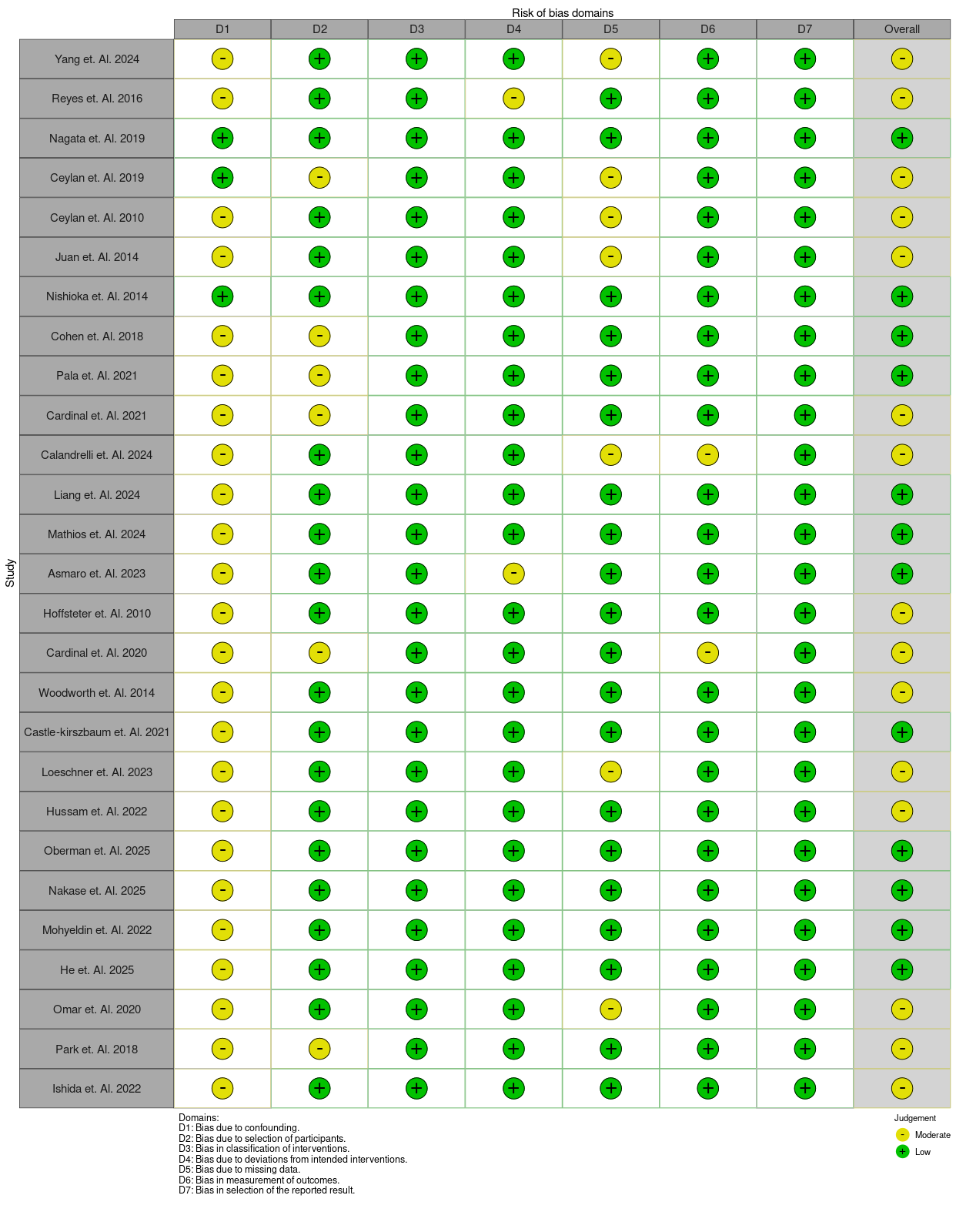
